# The Development of a Brain Injury Survivor Patient and Public Involvement Group by a Brain Injury Survivor

**DOI:** 10.1101/2024.04.14.24305787

**Authors:** James Piercy, Colin Hamilton, Robert Runcie, Christi Deaton, Alexis J Joannides

## Abstract

**Background:** Patient and public involvement (PPI) in research is seen as key to ensuring applicability and impact. Undertaking PPI in people after brain injury has long been seen to be a challenge. In 2020 The NIHR Brain Injury MedTech Cooperative developed a programme with the aim of improving PPI involvement, impact and diversity in this population.

**Methods:** Through a process of iterative development, a PPI programme was created. It built on an existing underutilised database of people after brain injury and their carers who were interested in engaging with PPI and utilised video-calling software. It was led by a Brain injury Survivor acting as Facilitator with admin support from the MedTech Cooperative.

**Results:** To date 14 PPI sessions were completed supporting a total of 17 projects. The diversity of the panel members was comparable to that of the population at large. However, further work is needed, especially in engaging people experiencing homelessness, people living outside of England and those with communication impairments. Feedback from researchers was positive and specific impacts are stated.

**Conclusion:** Through the leadership of a facilitator who has an understanding of the lived experience of brain injury a PPI programme has been developed. The use of a video-calling platform enabled a wider representation then a face-to-face group would have and techniques such as shortened sessions and single project presentations ensured engagement and impact.

## Background

Patient and public involvement (PPI) is acknowledged to be central to good research practice(Brett et al., 2014). However, enacting it in practice is a challenge(Brett et al., 2014). This can be particularly true in populations who may experience disability, with researchers finding it difficult to engage with these patients and their carers(Brett et al., 2014). These challenges can lead to inequities in access to PPI opportunities.

Populations who have limited access or particular difficulties to engaging with research are less likely to benefit from the improvements in care that research can bring(McDonald & Keys, 2008). Research is also likely to be less applicable and wasteful if rigorous PPI with disadvantaged groups is not conducted (Brett et al., 2014). Due to the emphasis that funders now place on PPI, research projects without robust PPI with relevant groups may find accessing funding more challenging, leading to further health inequalities in such populations.

Brain injury is a leading cause of death and disability worldwide(Dewan et al., 2019). With sequelae including physical impairments, cognitive deficits, and communication challenges(Turkstra, Politis, & Forsyth, 2015) brain injury survivors face many barriers to actively participating in PPI. Caring responsibilities may limit the ability for their family and carers to engage(Aitken et al., 2009). Therefore, finding effective methods of engaging with and facilitating research involvement among brain injury survivors are key to ensuring that research is relevant to this population(Whitehouse et al., 2021).

The NIHR Brain Injury MedTech Co-operative (Brain Injury MIC) works with patients, carers, NHS, charities, academia, inventors, small and medium sized enterprises (SME) and potential investors to support the development of new medical devices and healthcare technologies improving the effectiveness and quality of healthcare services. Since its inception the Brain Injury MIC has established a patient advisory group (PAG). This group is formed of those who have suffered a brain injury and carers of people who have had a brain injury. The PAG provides a governance role to the Brain Injury MIC as well as contributing to strategic direction. It has been key in a push to develop an effective PPI strategy. In this paper we describe the development of the Brain Injury MIC’s PPI programme, initiated by Brain Injury Survivors, led by a Brain Injury Survivor and contributed to by Brain Injury Survivors.

### Previous PPI Offering and Identification of Gap

Until 2020 PPI in research supported by the Brain Injury MIC was dealt with by the individual research teams with only informal help from the MIC administrative team. This support entailed advertising projects to members of the Register for Healthcare Involvement and Technology Evaluation (RHITE) database (www.brainmic.nihr.ac.uk/RHITE). RHITE was developed in 2012 by a previous version of the Brain Injury MIC and is a database of people with brain injury and their families/cares who were potentially interested in supporting research. While the RHITE could be a means of connecting researchers to interested parties it was underused.

Following discussion, the PAG and Brain Injury MIC administration agreed that this under-use of the RHITE resource was making it difficult to achieve meaningful and timely patient involvement in line with the UK standards for Public Involvement(Partnership, 2019). A more focussed support programme for PPI was needed, especially to assist researchers at application stage to improve their research questions in discussion with brain injury survivors and to demonstrate early commitment to PPI.

An outreach programme was proposed by the Brain Injury MIC with support of the PAG to make connections between those with lived experience of brain injury on the RHITE database and research groups developing new interventions on the acquired brain injury pathway.

#### Aims

The aims of the outreach programme were:

- To increase involvement of brain injury survivors and their family or carers in the development of brain injury research
- To widen the diversity of involved members to be more representative of the population as a whole. This to include but not be limited to geography, gender, age, ethnic group and educational background.

### Iterative Development

The outreach programme was developed through a process of discussion and consensus building with the Brain Injury MIC and the PAG. It was led by a brain injury survivor active in public engagement who took the role of Facilitator. Administrative and financial support was provided by the Brain injury MIC. The Outreach Programme adopted a model of iterative evaluation and review after initial launch and subsequent sessions.

The initial meeting between the Brain Injury MIC and the Facilitator was undertaken in Nov 2020 to set expectations for the programme and format. Due to the Covid-19 pandemic the meeting participants agreed that an online platform was needed to protect patients, carers and researchers. The participants also discussed their expectations of what the programme in the first year should deliver:

- A series of online events suitable for patients and carers, in which they can hear about research being supported by the Brain Injury MIC and discuss with the principal investigator.
- A supporting document for people joining the sessions, with links to the Brain Injury MIC, its research programmes and ways to get involved in research.
- Three outreach sessions within 6 months
- A review of the outreach programme after the first three sessions

#### Pilot stage: (Month 0-3)

A survey was sent to the investigators of all supported research projects to gather information about the researchers’ requirements for involvement and participation of patients, carers and the public in their projects, for example, review of documents or serving on an advisory committee (appendix 1). Each group was asked to provide a lay summary of their project. Six completed forms were initially received and from these 3 were chosen to be featured in the outreach session.

A pilot event was set up to test the effectiveness of the model on attendance rates of PPI Panel Members and impacts on projects. It was run using the GoTo Meeting™ web platform. Potential Panel Members were identified through the RHITE database and by contact with the local Headway (www.headway.org.uk) group which offers support and signposting for brain injury survivors and their families. Invitations were sent to those registered on the RHITE database and through Headway to its contact list. The event was set to run for 90 minutes and featured an introduction to the Brain Injury MIC followed by a presentation on the difference between engagement, involvement and participation.

Examples of current projects were discussed by the Brain Injury MIC to demonstrate the differences between engagement, involvement and participation as well as highlight opportunities to be engaged. Panel Members were invited to register their interest in specific projects by contacting the programme manager or to register a more general interest in research by joining the RHITE database.

In line with best practice and following the guidelines from the Centre for Engagement and Dissemination(NIHR, 2022) participants were offered reimbursement in the form of a £25 voucher. Time was allowed for questions and comments, with participants invited to share their own experiences of involvement in research. After the session all delegates were sent a copy of the outreach brochure (Appendix 2) and reminded of the links to join RHITE. Contact information was provided to give feedback and ask further questions.

#### Stage 1: (Month 4-7)

Following the pilot session, a team discussion took place incorporating comments from Panel Members and reflection from the Facilitator. The following changes were made:

- Reduced duration
- Switch to Zoom. The platform was felt to be more versatile, and attendees were more familiar with using it.
- Shorter introduction. The overview of the Brain Injury MIC activity was reduced to a very short introduction.
- Less time given to education on the distinctions between participation, involvement and engagement.
- Guest speaker. An aim was set to invite a researcher to each outreach session to speak personally about their project.
- Improved web visibility. The Brain Injury MIC website was updated to include information about the programme, engagement, involvement and participation and how to access the RHITE database.

Three research projects were featured in each of the 4 online workshops in stage 1. One of these was given prominence with the inclusion of a presentation from the research team, but due to time constraints the other two were presented by the Facilitator. The researchers could ask the group direct questions about their research question, methodology and recruitment strategy. With consent from the Panel members, sessions were recorded and shared with the research team after the event.

#### Stage 2: (Month 8-20)

Feedback from researchers and Panel Members indicated that whilst featuring a number of research projects allowed for variety and wider interest, a more focussed approach might yield greater impact. Delegates told the team that duration should be limited to 1 hour and that morning slots were easier to handle. Delegates emphasised that the cognitive fatigue experienced by many brain injury survivors made sustained attention difficult.

In the most recent iteration, a single project was featured presented by a member of the research team. The sessions were facilitated by the Facilitator. Key questions and issues which could be informed by the lived experience of brain injury survivors and their carers were agreed before the focus groups and time was allotted to cover these.

Greater attention was paid to recruitment of PPI Panel Members with efforts made to match people with direct experience of the topic under discussion and to improve the geographic, gender, age and ethnic diversity of the group. As well as existing contacts the events were publicised through the People in Research website (https://www.peopleinresearch.org). Interested parties were invited to apply and their suitability explored with simple questions about the nature of their direct experience, location and ethnic background.

To gain an understanding of progress against the stated aims of this project (Increasing involvement in PPI and increasing the diversity of PPI Panel Members) numbers of Panel Members attending each session were recorded and a demographic questionnaire was sent out to gain an understanding of the makeup of the group. To assess impacts of the PPI group on each research study presented, feedback forms were issued to all investigators of projects that were supported by the outreach programme. The forms were based on the Centre for Research in Public Health and Community Care Guidance for Researchers: Feedback document (Mathie, 2018).

## Results

Since the start of the PPI group 17 projects have been presented. This included 3 from Small and Medium Sized Enterprises (SME). Eight were featured in the initial outreach document and presented in stage 1, a further nine presented by researchers in the sessions held during stage 2 (including the 3 projects from SMEs). A timeline and attendance at the outreach sessions are presented in Table 1.

**Table 1:** Number of Delegates Attending Each PPI session.

|  | **Month since start of project** | **Delegates Registered** | **Delegates Attending** |
| --- | --- | --- | --- |
| Pilot session | Month 3 | 16 | 11 |
| Stage 1 sessions | Month 4 | 18 | 10 |
|  | Month 5 | 10 | 6 |
|  | Month 6 | 5 | 0 |
|  | Month 7 | 8 | 8 |
| Stage 2 sessions | Month 11 | 15 | 13 |
|  | Month 12 | 9 | 9 |
|  | Month 13 | 19 | 12 |
|  | Month 14 | 9 | 7 |
|  | Month 16 | 27 | 22 |
|  | Month 17 | 14 | 9 |
|  | Month 18 | 11 | 11 |
|  | Month 18 | 13 | 13 |
|  | Month 20 | 10 | 7 |

The demographic makeup of the overall group can be found in Table 2 with geographical location in Figure 1.

**Table 2.**
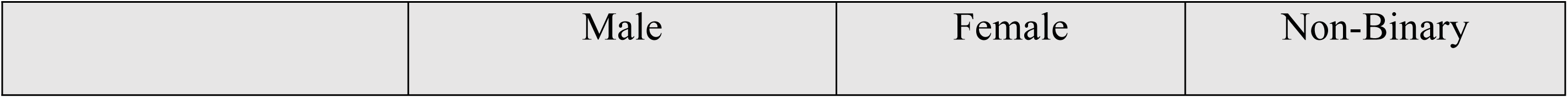
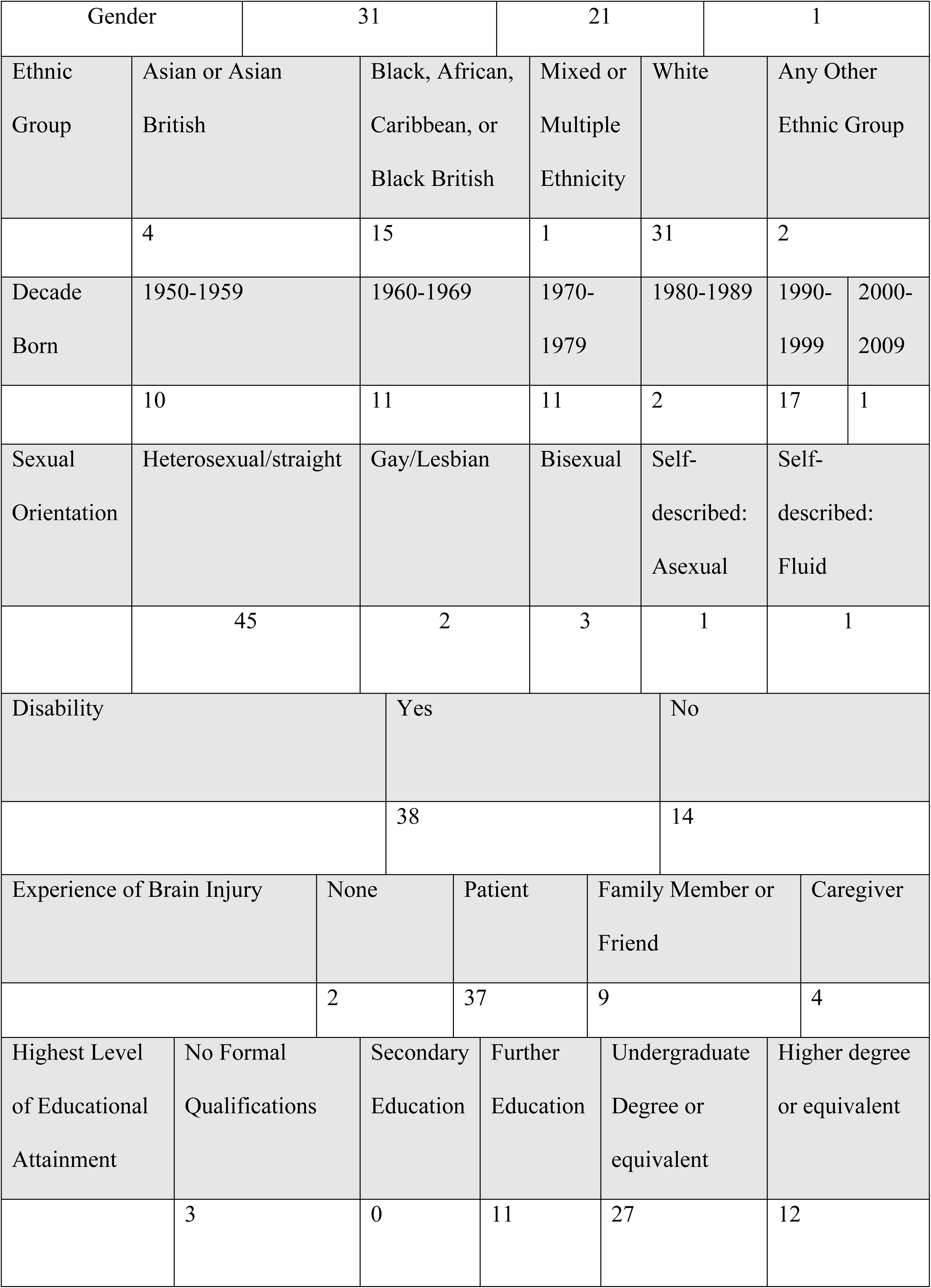
Demographics of the PPI Group.

|  | Male | Female | Non-Binary |
| --- | --- | --- | --- |

|  |  |  |  |  |  |  |
| --- | --- | --- | --- | --- | --- | --- |
| Gender |  | 31 |  | 21 |  | 1 |
| Ethnic Group | Asian or Asian British | Black, African, Caribbean, or Black British | Mixed or Multiple Ethnicity | White | Any Other Ethnic Group |  |
|  | 4 | 15 | 1 | 31 | 2 |  |
| Decade Born | 1950-1959 | 1960-1969 | 1970-1979 | 1980-1989 | 1990-1999 | 2000-2009 |
|  | 10 | 11 | 11 | 2 | 17 | 1 |
| Sexual Orientation | Heterosexual/straight | Gay/Lesbian | Bisexual | Self-described: Asexual | Self-described: Fluid |  |
|  | 45 | 2 | 3 | 1 | 1 |  |
| Disability |  | Yes |  |  | No |  |
|  |  | 38 |  |  | 14 |  |
| Experience of Brain Injury |  | None | Patient | Family Member or Friend |  | Caregiver |
|  |  | 2 | 37 | 9 |  | 4 |
| Highest Level of Educational Attainment | No Formal Qualifications | Secondary Education | Further Education | Undergraduate Degree or equivalent |  | Higher degree or equivalent |
|  | 3 | 0 | 11 | 27 |  | 12 |

**Figure 1.**
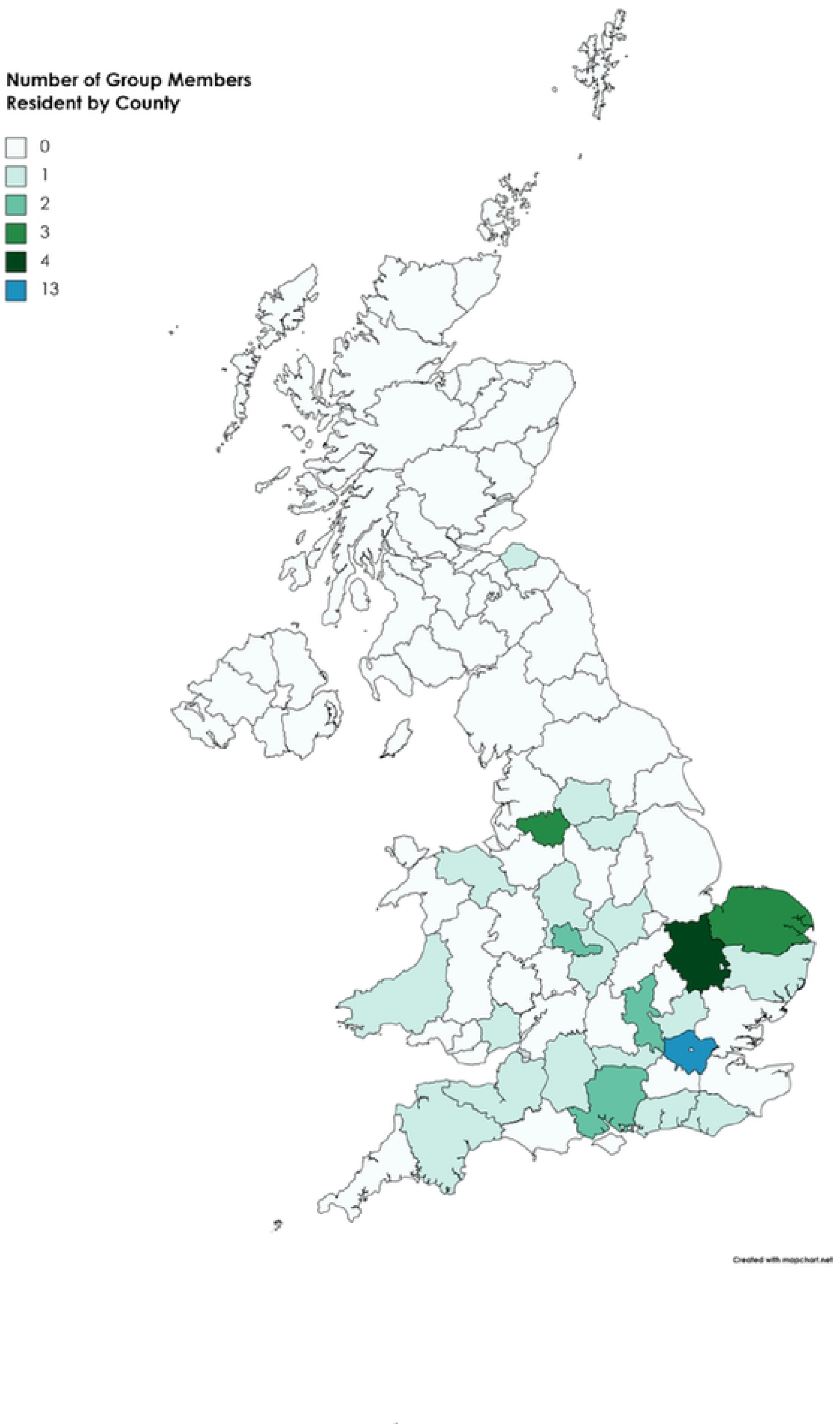
Geographical Distribution of PPI Group Members by County.

Out of the 17 projects supported only investigators from seven projects provided feedback when requested. Results can be found in table 3. While numbers are small all projects found the group “Very Helpful” or “Quite Helpful” and all investigators that responded to the question (n=6) would recommend the group to colleagues. In two projects no changes were made with one discussing that concerns were raised by the PPI group about data collection, but further clarification was sought, and no changes were needed. While not a change, this may have strengthened the proposal and confirmed that the approach taken was acceptable. Comments were received on the facilitation of the session which helped with its development and improvement.

**Table 3:**
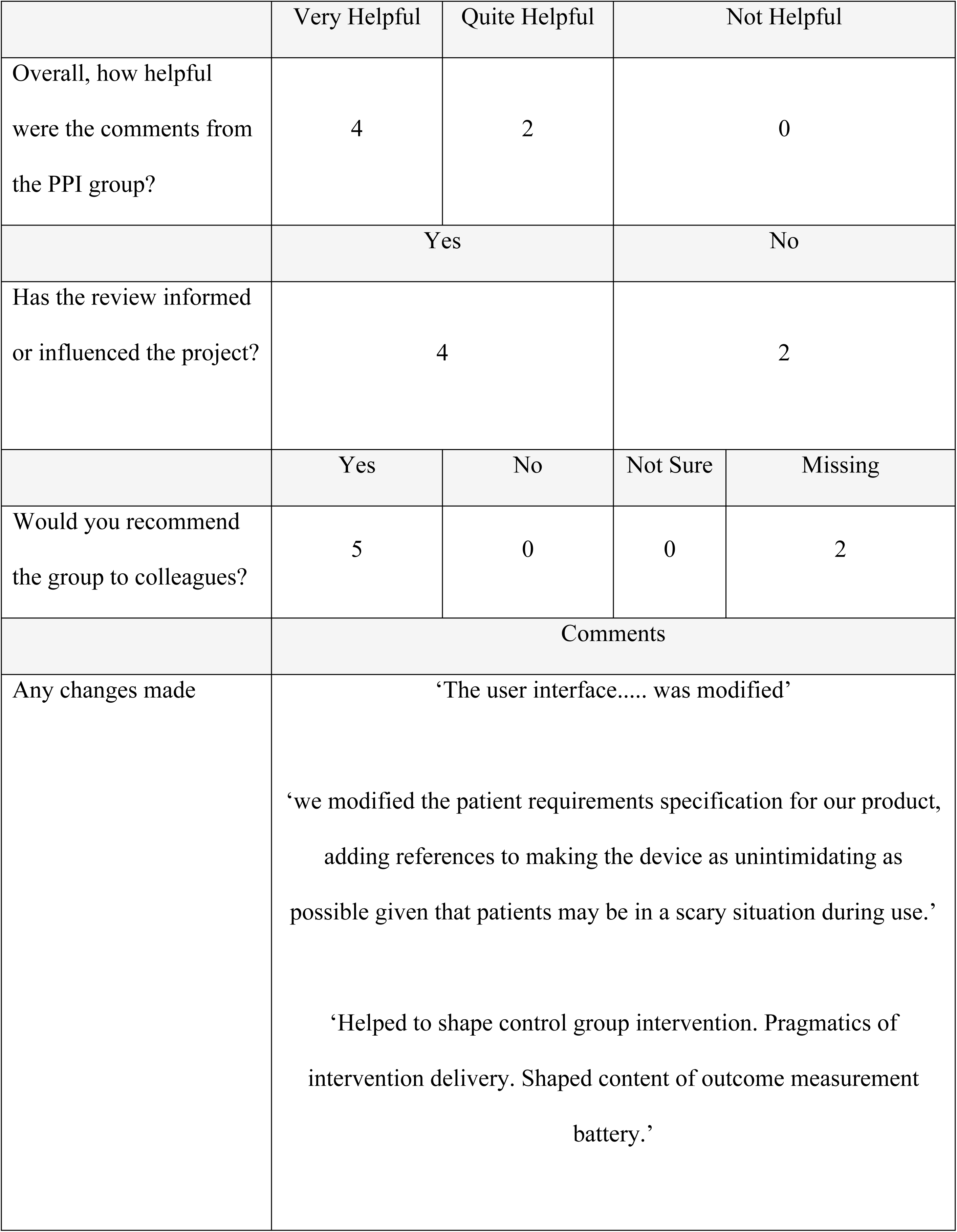
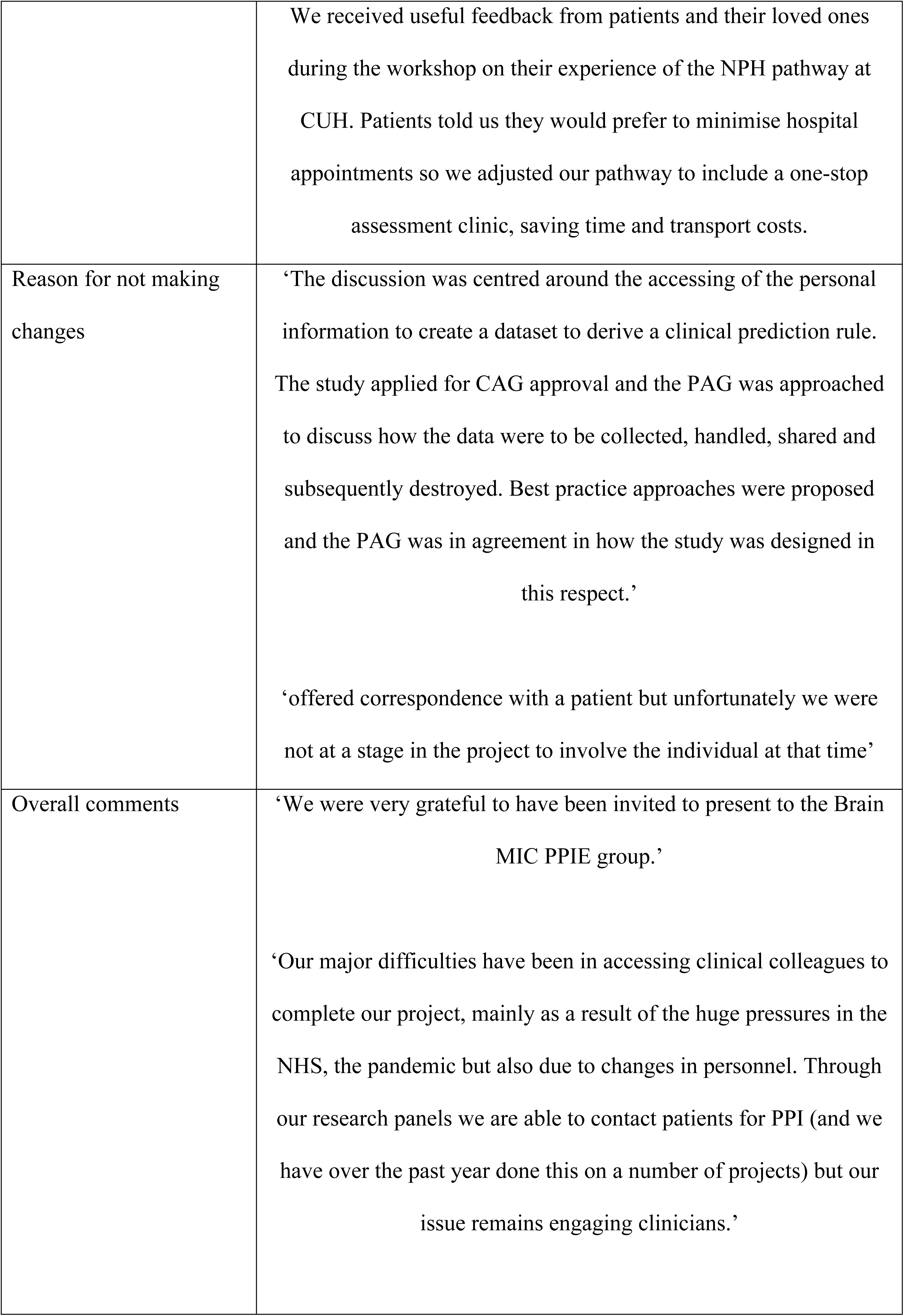
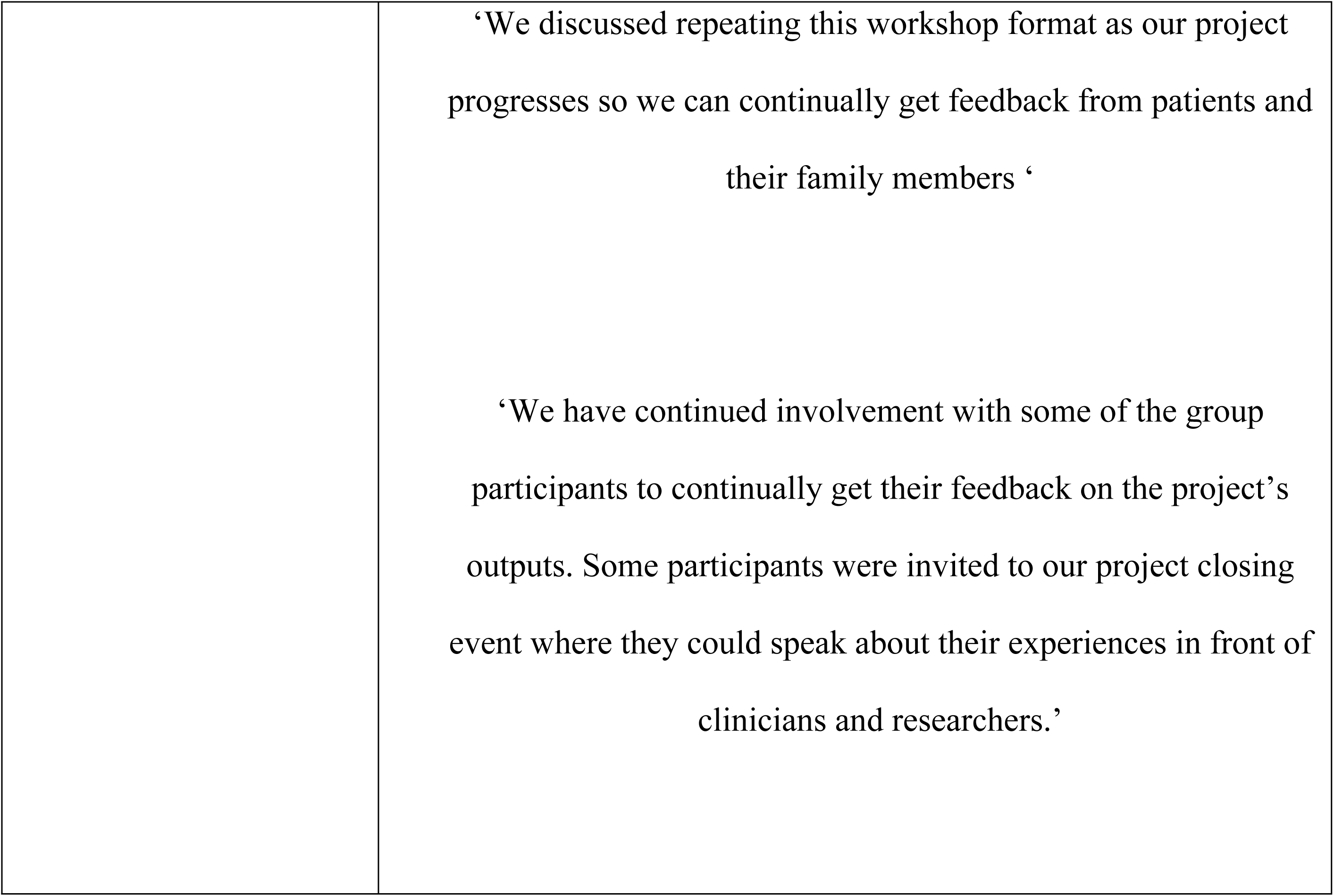
Feedback from Researchers.

“We had a very positive experience in part because we were able to send private messages to the facilitator during the session to keep the conversation on track. The facilitator said they would adopt this for future sessions. This worked particularly well with it being a virtual format as it would be harder to communicate privately in person and not disrupt the conversation.”

## Discussion

The development of meaningful PPI is often stated as a challenge within research(Ocloo, Garfield, Franklin, & Dawson, 2021). Various methods and models have been described in the literature(South et al., 2016) however this example emphasises the flexibility advocated by Buck et al (Buck et al., 2014). From the start the team adopted a process driven by the PPI Panel Members of iterative evaluation, reviewing the format and potential impact after each session to deliver the aims of the project. Overall, the programme was successful in the stated aims of facilitating a diverse group of people after brain injury to participate in impactful PPI activities. However, work remains to be done. Key points are presented below.

### Understanding the Population

Brain Injury can cause a wide range of issues and understanding the challenges faced by survivors and their families or carers was key to the development of this project. From the initial idea the PAG, formed of people who have suffered brain injuries and their carers, has been key in ensuring the project was of importance to this population. This was further developed by the Facilitator who is a survivor of severe brain injury and whose direct experience helped inform this process and was key to the success.

### Ensuring Impact

The aim of the outreach project was to have real involvement from Panel Members, listening to their thoughts and responses to issues more than disseminating information about projects. It aimed to have useful input from lived experience at all stages of the research cycle. Informing preapplication strategy, focussing research questions and modifying delivery of research projects which fit well with the NIHR UK Standards for Public Involvement (7). The importance of being able to meaningfully input on research projects has been described before (11) and is seen to be a key element of PPI. Through the iterative development the Panel moved from aiming to influence a larger number of projects with less input from researchers to intensely inputting on one in each session. This enabled active input rather than a light touch approach, which the group found to be key in ensuring impact.

The role of an active and experienced Facilitator was found to be vital in ensuring that messages were understood and that the purpose of the session was clear to participants. Prior to each session the facilitator reviewed the researchers’ presentations to check for clarity, and in a pre-meeting, identified key questions or areas for discussion with the research team. The use of plain English was mandatory at all points. In the Panel Meeting the facilitator directed the questions and ensured Panel Members understanding and supported their participation to maximise impact.

### Maximising Accessibility

**FIGURE 2.**
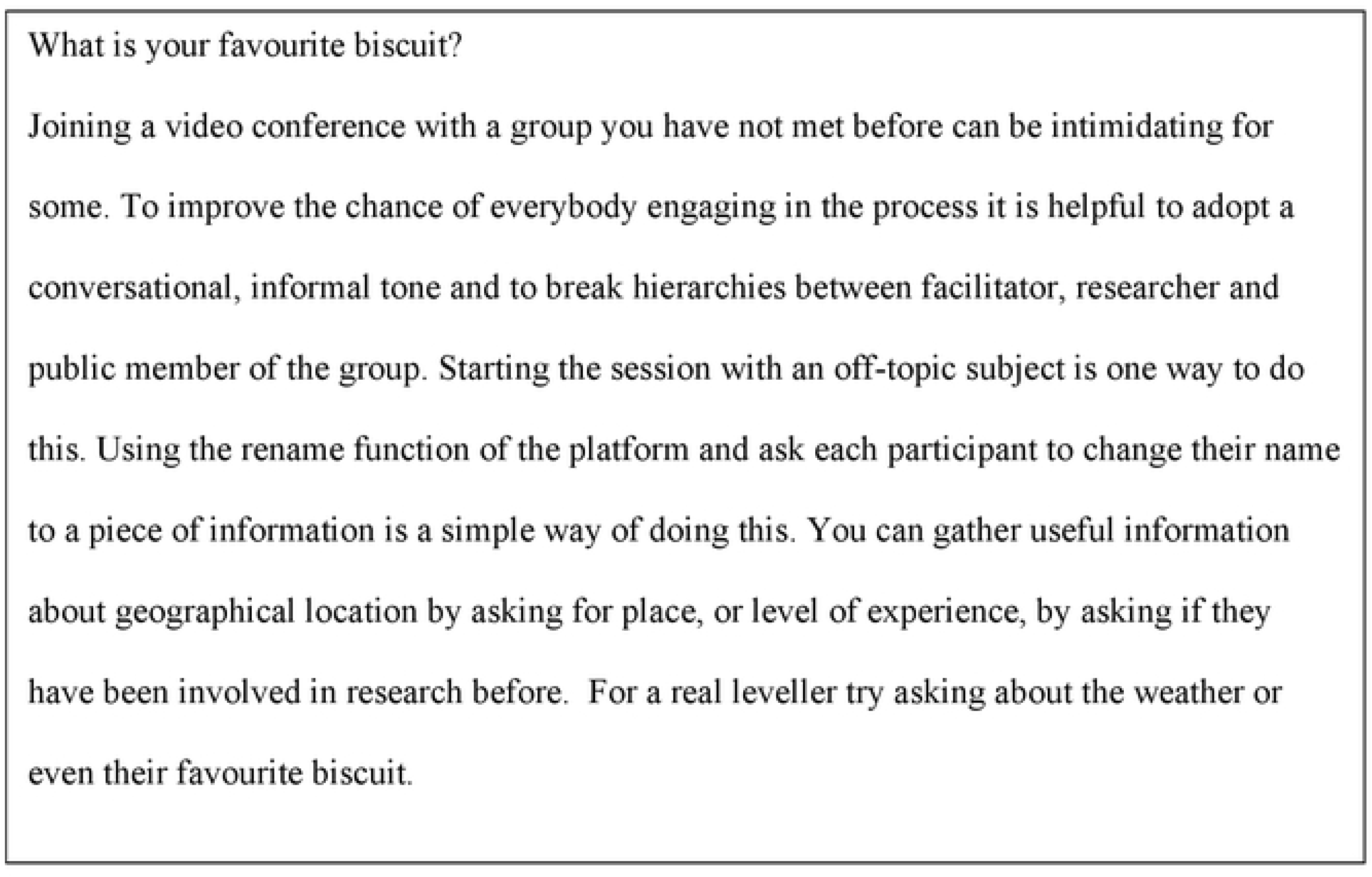
Ice-Breakers for Online Groups.

By understanding the population, a focus on accessibility was acknowledged to be central in order to optimise participation. While the choice of an online process was initially driven by the COVID pandemic, which made face-to face sessions impossible, the need to engage with vulnerable people meant that in-person group meetings were not feasible. The online format has worked well as a means of engaging patients and the public with research projects, allowing people from across the UK to take part in the process and, whilst there is still work to do, it appears to have widened the diversity of participants. The use of the Zoom platform to host the events was driven by the familiarity of the system to users. The pandemic also meant that potential participants were likely to be familiar with the technology and able to engage without prolonged instruction. The widespread use of online platforms in PPI is a relatively new phenomenon (12) and the advantages and disadvantages are still being evaluated. It must be acknowledged, however, that there are groups that find accessing technology more difficult and by using this platform certain groups are may have been disadvantaged(Das & Gonzalez, 2020).

There are particular issues of importance for engaging people in PPIE after brain injury. Brain injury survivors commonly live with issues caused by cognitive fatigue and screen-based activity has been shown to be more likely to cause these issues (Bailenson, 2021), which can limit participation. In response to these concerns sessions have been time limited to 1 hour and following feedback from participants have been scheduled before lunch rather than in the afternoon when people are more likely to be tired. The group format also means that people with severe communication impairment are more likely to find the sessions difficult to engage with(Paterson & Carpenter, 2015). The team is looking into offering bespoke sessions in these cases (Palmer & Paterson, 2013).

Researchers found the inbuilt ability in the Zoom platform to record and review the session at a later date valuable. This allowed follow up questions to be directed to participants and a good understanding of contentious areas in discussion. The use of ‘chat’ functions allowed participants to note questions as they think of them, rather than having to remember them and finding a time to raise them later, which in view of the high incidence of memory difficulties in this population(Paterno, Folweiler, & Cohen, 2017) can be a particular challenge. The facilitators also found that these questions could be used to restart conversations or to redirect conversation which may have become fixed on a minor point.

It is well acknowledged that PPI group members should be reimbursed for their time and discussion(Manikandan et al., 2022). Legislation around HMRC IR35 can make it problematic for public bodies to make direct payments to individuals so provision of vouchers for payment was selected, which has been found to be effective previously(Manikandan et al., 2022). Contributors reported being happy with this model. Research programmes at application stage do not have funding to support PPI but the Research Design Service public involvement fund was able to cover the cost of these initial sessions.

### Ensuring Representation

Diversity in PPI is an acknowledged issue (INVOLVE, (2012)). The project aimed to improve the geographic, ethnic and socio-economic diversity of the group and the final results showed some success. An initial challenge of the team was not having previous data on the diversity of the initial participants in the RHITE database. The group so far is England centric with a concentration in London which does limit range of views. With other factors, however the group was more representative of the country with 10% of the PPI group identifying as LGBT+ in comparison with 10.6% in the most recent census(Statistics, 2022). With 28% of the group describing themselves as Black, African, Caribbean, or Black British this group is a higher proportion then in the wider population (4% (Statistics, 2022)). One person stated that they were “homeless” which is often seen to be one of the most challenging populations to engage with PPI work (Dawes, Barron, & Lee, 2022). People experiencing homelessness however are at significantly higher risk for ill health and brain injury with 53.1% suffering a traumatic brain injury across their lifetime(Stubbs et al., 2020) and expanding input from this population is likely to provide significant new insights for the group.

The wide ranges of presentations that people have after Brain Injury (Turkstra et al., 2015) also make ensuring appropriate representation difficult. At present no attempt is made to understand diversity of impairment within the group but is seen to be a key future development.

### Moving forward

The latest review of the programme has identified areas where the outreach session could be improved further.

- More effort needs to be made to improve the diversity of those attending the sessions. Identifying third sector organisations and support groups for communities of ABI survivors with specific types of injuries or conditions might be one way of doing this. In particular, geographical diversity will need to be further addressed.
- A related project to develop a patient portal is underway. This will improve public access to information about research projects, allowing greater transparency of the work of the Brain Injury MIC and act as a forum to recruit study participants and Panel Members.
- Currently the PPI group has been focused on early-stage projects. As the supported projects continue and move to stages such as dissemination the PPI group will have input and impact on these aspects.
- Further focussing on increasing awareness of the use of the PPI programme with SMEs and supporting them to develop solutions and products to address problems faced by Brain Injury Survivors.

## Conclusion

We have illustrated the development of an outreach programme, led by a Brain Injury Survivor that engages brain injury survivors and their families or carers in learning about and providing input to research being conducted by the Brain Injury MIC. The programme has grown and been able to engage relevant and increasingly diverse participants across much of the UK. More researchers in brain injury are engaging with the programme and finding it useful for their research. While the Brain Injury MIC team started as novices in this area, through an iterative process a purposeful and useful programme was formed which has had an impact on research within an area that is often challenging in which to obtain patient and carer input. Key learning from this programme includes ensuring measurement of diversity, supporting access through the use of online platforms, maximising impact through preparation of researchers ahead of the sessions, and a knowledgeable and empathetic facilitator.

## Data Availability

Data cannot be shared publicly because of data protection rules. Data are available from the the NIHR Brain Injury Med Tech Cooperative on request by emailing

## List of abbreviations

ABI: Acquired Brain Injury
MIC: Med tech cooperative
PAG: Patient advisory group
PPI: Patient public involvement
PPIE: Patient public involvement and engagement
SME: Small and medium enterprise

## Declarations

### Ethics approval and consent to participate

Not applicable

### Consent for publication

All authors consent to publication

### Availability of data and materials

All material is owned by the authors and available on request

## Acknowledgements

The Authors wish to thank the members of the Brain Injury MIC PAG as well members of RHITE and all those who have attended as a PPI Panel member.

The research was supported by the National Institute for Health and Care Research (NIHR) Brain Injury MedTech Co-operative based at Cambridge University Hospitals NHS Foundation Trust and University of Cambridge. The views expressed are those of the author(s) and not necessarily those of the NHS, the NIHR or the Department of Health and Social Care.

## Declarations

### Acknowledgements

The authors would like to thank all of the patients and carers for their contribution to then outreach project and ongoing support of research

## References

Aitken, M. E., McCarthy, M. L., Slomine, B. S., Ding, R., Durbin, D. R., Jaffe, K. M., Mackenzie, E. J. (2009). Family burden after traumatic brain injury in children. Pediatrics, 123(1), 199–206. doi:10.1542/peds.2008-0607

Bailenson, J. N. (2021). Nonverbal overload: A theoretical argument for the causes of Zoom fatigue. Technology, Mind, and Behavior, 2(1). doi:10.1037/tmb0000030

Brett, J., Staniszewska, S., Mockford, C., Herron-Marx, S., Hughes, J., Tysall, C., & Suleman, R. (2014). Mapping the impact of patient and public involvement on health and social care research: a systematic review. Health Expectations, 17(5), 637–650. 10.1111/j.1369-7625.2012.00795.x

Buck, D., Gamble, C., Dudley, L., Preston, J., Hanley, B., Williamson, P. R., & Young, B. (2014). From plans to actions in patient and public involvement: qualitative study of documented plans and the accounts of researchers and patients sampled from a cohort of clinical trials. BMJ Open, 4(12), e006400. doi:10.1136/bmjopen-2014-006400

Das, L. T., & Gonzalez, C. J. (2020). Preparing Telemedicine for the Frontlines of Healthcare Equity. Journal of General Internal Medicine, 35(8), 2443–2444. doi:10.1007/s11606-020-05941-9

Dawes, J., Barron, D. S., & Lee, L. E. (2022). Capturing learning from public involvement with people experiencing homelessness to help shape new physiotherapy research: Utilizing a reflective model with an under-served, vulnerable population. Health Expectations, 25(5), 2203–2212. 10.1111/hex.13397

Dewan, M. C., Rattani, A., Gupta, S., Baticulon, R. E., Hung, Y.-C., Punchak, M., Park, K. B. (2019). Estimating the global incidence of traumatic brain injury. Journal of Neurosurgery, 130(4), 1080–1097. doi:10.3171/2017.10.jns17352

INVOLVE. (2012). Strategies for diversity and inclusion in public involvement: Supplement to the briefing notes for researchers. INVOLVE, Eastleigh. Retrieved from https://www.invo.org.uk/wp-content/uploads/2012/06/INVOLVEInclusionSupplement1.pdf

Manikandan, M., Foley, K., Gough, J., Harrington, S., Wall, É., Weldon, F., Fortune, J. (2022). Public and Patient Involvement in Doctoral Research During the COVID-19 Pandemic: Reflections on the Process, Challenges, Impact and Experiences From the Perspectives of Adults With Cerebral Palsy and the Doctoral Researcher. Frontiers in Rehabilitation Sciences, 3. doi:10.3389/fresc.2022.874012

Mathie, E. (2018). Guidence for Researchers: Feedback. Retrieved from https://www.clahrc-eoe.nihr.ac.uk/wp-content/uploads/2016/05/Guidance-for-Researchers-PPI-Feedback_2018.pdf:

McDonald, K. E., & Keys, C. B. (2008). How the powerful decide: access to research participation by those at the margins. Am J Community Psychol, 42(1-2), 79–93. doi:10.1007/s10464-008-9192-x

NIHR. (2022). Payment guidance for researchers and professionals. Retrieved from www.nihr.ac.uk/documents/payment-guidance-for-researchers-and-professionals/27392

Ocloo, J., Garfield, S., Franklin, B. D., & Dawson, S. (2021). Exploring the theory, barriers and enablers for patient and public involvement across health, social care and patient safety: a systematic review of reviews. Health Research Policy and Systems, 19(1), 8. doi:10.1186/s12961-020-00644-3

Palmer, R., & Paterson, G. (2013). To what extent can people with communication difficulties contribute to health research? Nurse Res, 20(3), 12–16. doi:10.7748/nr2013.01.20.3.12.c9491

UK Public Involvement Standards Development Partnership (2019) UK Standards for Public Involvement. Retrieved From www.nihr.ac.uk/pi-standards/standards

Paterno, R., Folweiler, K. A., & Cohen, A. S. (2017). Pathophysiology and Treatment of Memory Dysfunction After Traumatic Brain Injury. Curr Neurol Neurosci Rep, 17(7), 52. doi:10.1007/s11910-017-0762-x

Paterson, H., & Carpenter, C. (2015). Using different methods to communicate: how adults with severe acquired communication difficulties make decisions about the communication methods they use and how they experience them. Disability and Rehabilitation, 37(17), 1522–1530. doi:10.3109/09638288.2015.1052575

South, A., Hanley, B., Gafos, M., Cromarty, B., Stephens, R., Sturgeon, K., Vale, C. L. (2016). Models and impact of patient and public involvement in studies carried out by the Medical Research Council Clinical Trials Unit at University College London: findings from ten case studies. Trials, 17, 376. doi:10.1186/s13063-016-1488-9

UK Census (2021). Office of National Statistics, Available at https://census.gov.uk/census-2021-results

Stubbs, J. L., Thornton, A. E., Sevick, J. M., Silverberg, N. D., Barr, A. M., Honer, W. G., & Panenka, W. J. (2020). Traumatic brain injury in homeless and marginally housed individuals: a systematic review and meta-analysis. The Lancet Public Health, 5(1), e19–e32. doi:10.1016/S2468-2667(19)30188-4

Turkstra, L. S., Politis, A. M., & Forsyth, R. (2015). Cognitive–communication disorders in children with traumatic brain injury. Developmental Medicine & Child Neurology, 57(3), 217–222. 10.1111/dmcn.12600

Whitehouse, D., Piffer, F., Becker, T., Gravett, K., Stewart, A., Basi, K., Newcombe, V. (2021). Challenges, approaches and opportunities for Patient and Public Involvement (PPI) in Traumatic Brain Injury (TBI) research. Br J Neurosurg, 35(5), 651–652. doi:10.1080/02688697.2021.1922605

